# Peripheral Blood Mononuclear Cell-Derived miR-664a-3p is Associated with Plaque Burden and Necrotic Core Characteristics in Coronary Artery Disease Across Two Independent Populations

**DOI:** 10.64898/2026.06.23.26355128

**Authors:** Bhanu Duggal, Atul K. Kashyap, Ghanshyam Kumar, Sathyamangla V. Naga Prasad

## Abstract

**Background:** Stability of the atherosclerotic plaque in coronary artery disease (CAD) is determined by features such as total plaque burden and necrotic core volume. Since invasive procedures are required to evaluate plaque stability, we tested whether the peripheral blood mononuclear cell (PBMC) microRNA (miR) signature could correlate with measures of plaque stability and thus serve as a non-invasive biomarker.

**Method:** Patients from two distinct geographical locations in India were recruited to the study (Site 1: CAD=19, non-CAD=5; Site 2: CAD=12, non-CAD=7) and underwent invasive intravascular ultrasound with virtual histology to assess plaque burden and necrotic core volume. RNA from PBMCs of these patients was subjected to unbiased sequencing. Differential miR expression evaluated by DESeq2 and assessed for co-relationship with plaque stability. miR target gene prediction was performed using multiple databases, and Enrichr was used for enrichment analysis.

**Results:** Unbiased RNA sequencing identified miR-664a-3p to be significantly downregulated in CAD patients from both sites (Site 1: log2FC=−1.02, p=0.0033 & Site 2: log2FC=−1.04, p=0.0007). miR-664a-3p expression was inversely correlated with plaque burden and necrotic core volume. Receiver operating characteristic (ROC) analysis of miR-664a-3p showed significant discriminative performance in the CAD cohort, with AUC values of 0.842 (Site 1) and 0.881 (Site 2). miR664a-3p target prediction and pathway enrichment analysis revealed selective enrichment of inflammatory signaling pathways, such as IL-17 and TNF, suggesting an association between PBMC pro-inflammatory response and plaque vulnerability.

**Conclusion:** miR-664a-3p is downregulated in CAD patients and inversely correlates with measures of plaque stability, with potential as a biomarker for identifying patients at risk of CAD progression and plaque instability.

## Introduction

Coronary artery disease (CAD) causes a substantial global health burden, with ∼422 million prevalent cases of CAD worldwide, with the largest contributor being the South Asian Region (1). This is supported by the evidence that the age-standardized prevalence of CAD in South Asia is 2393 per 100,000 (95% uncertainty interval: 1746–3188), underscoring a substantial regional burden of disease (2,3). Furthermore, the INTERHEART study reported a lower mean age at first myocardial infarction among South Asians (∼53 years) compared with other countries (∼58.8 years) (4). Given India’s population, it is a significant contributor to the CAD death and mortality in the South Asian Region. The GBD study finds that there is a 2.3-fold increase in the prevalence of CAD (ischemic heart disease) in Indians in more recent times (5). This is further complicated by the observation that Indians present with cardiovascular diseases a decade earlier than their Caucasian Counterparts (5). Therefore, early identification and precise characterization of CAD in the Indian population are important to potentially develop proactive treatment strategies that prevent progression, reduce major adverse cardiovascular events (MACE), and lower cardiovascular mortality.

CAD is characterized by the progressive accumulation of atherosclerotic plaques within the arterial wall, which can lead to luminal narrowing and ischemia (6,7). While luminal stenosis is a primary diagnostic criterion, the plaque composition, specifically the presence of a lipid-rich necrotic core (NC) and a thin fibrous cap, is a critical determinant of lesion vulnerability and propensity to rupture (8). Patients with untreated lipid-rich coronary lesions, characterized by a high lipid core burden index, have been shown to carry a higher risk of future non-culprit lesion-related MACE. These findings highlight the clinical importance of identifying biomarkers that can detect high-risk, lipid-rich plaques (9) in Indians.

Although multiple studies have used circulating microRNAs (miRs) from plasma as potential surrogate markers for CAD (10,11), they may not have sufficient sensitivity to determine CAD progression or assess plaque stability. This is because plasma miRs could come from unrelated cells due to non-specific elevation of pro-fibrotic responses associated with pathology. Furthermore, it is difficult to determine the cells from which these miRs originate and whether they are secreted or released, given that cell death associated with pathology-associated pro-fibrotic signaling occurs. Such a response could skew the unbiased miR signature in plasma and may not consistently reflect the changes in plaque stability. To overcome these drawbacks, we have opted to use the peripheral blood mononuclear cell (PBMC) miR expression signature as a potential readout to reflect CAD progression and plaque stability. Use of PBMCs in our studies is based on the rationale that these cells dynamically respond to lesions, changes in luminal stenosis, or myocardial injury through an inflammatory response. Although these are localized changes, PBMCs undergo rapid transcriptional adaptations, including miR alterations, that are integral to mediating the inflammatory repair response and could serve as a consistent non-invasive biomarker. PBMCs serve as a source of biological information, providing chronological insights into disease characterization and exhibiting distinct inflammatory gene expression profiles in CAD patients (12,13). Since PBMCs mediate inflammatory responses that drive the progression of coronary atherosclerosis, analysis of miRs within their cellular compartment may provide greater pathobiological specificity.

miRs represent a class of small noncoding RNAs typically 18-24 nucleotides in length that regulate nearly 60% of human protein-coding genes by binding to the 3’ untranslated region of target mRNAs (9). Evidence indicates that miR expression is altered dynamically in response to pathophysiological stress. Our studies use an unbiased miR screening approach to identify a miR signature in PBMCs from CAD patients from India. These studies are pivotal, as very little is known about the PBMC miR signature in CAD patients of Indian ethnicity. This has significant clinical implications, given that the Indian population has a clear predisposition to CAD, and identifying non-invasive markers that may predict plaque stability can improve diagnosis and treatment.

## Methods

### Patient Population

We recruited two independent cohorts of participants from two distinct geographical locations in India. The first cohort was from Grant Medical College & Sir JJ Group of Hospitals, Mumbai (**Site 1**). Cases and controls were classified based on plaque burden. Twenty-four participants recruited at this site comprised of 19 CAD and 5 non-CAD patients. The second cohort was from the All-India Institute of Medical Sciences (AIIMS), Rishikesh, in North India (**Site 2**), with a rural patient population. Nineteen participants recruited at this site comprised of 12 CAD and 7 non-CAD patients. The study involved 43 participants, and the study design is depicted in **Figure 1**. The study was approved by the Institutional Ethics Committees at both tertiary care centers and hospitals, and written informed consent was obtained from all participants before enrolment.

**Figure 1:**
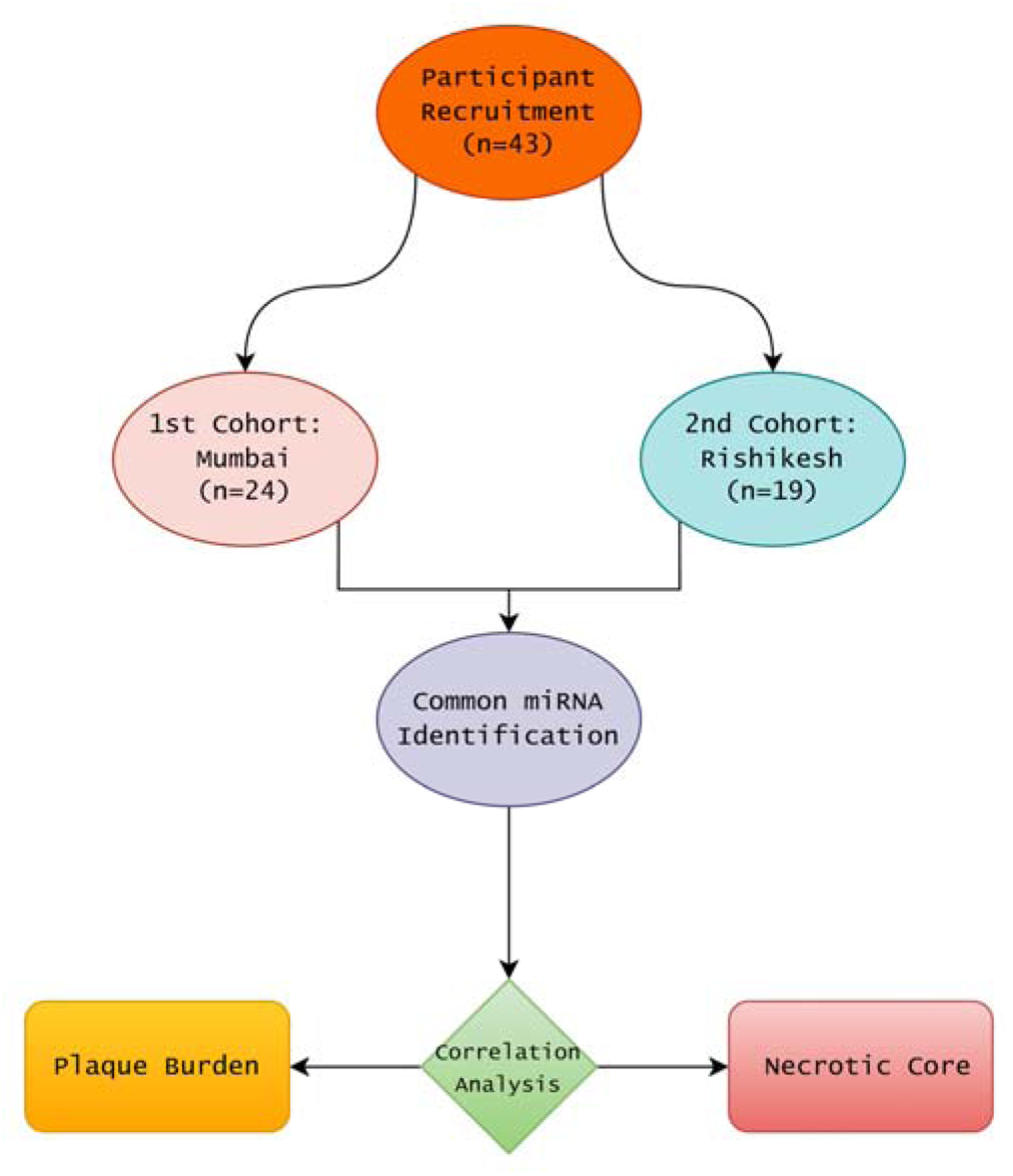
Overall Study Design.

### Intravascular ultrasound analysis (IVUS)

Participants for these studies were recruited from the cardiology outpatient department (OPD) who presented with angina and were undergoing coronary angiography. Patients accompanied by other medical complications such as cardiogenic shock, drug abuse, or arrhythmia were excluded from the study, and they represented a large population of patients undergoing IVUS at these centers. Participants recruited at both centers underwent IVUS imaging following intracoronary administration of 250 μg nitroglycerin using the Eagle Eye® Gold Catheter (20 MHz) at a pullback velocity of 1 mm/s. Both greyscale and virtual histology (VH-IVUS) imaging were performed, and raw radiofrequency data were captured at the peak of the R wave during ECG-gated VH-IVUS acquisition. The imaging data were archived on a CD, and volumetric analysis was performed using QIVUS (Research Edition 3.1, Medis Medical Imaging Systems, Leiden, The Netherlands). Offline analysis was performed by an independent investigator blinded to the clinical and angiographic data, followed by cross-verification by another investigator. After manual annotation of the lumen and vessel boundaries, the software automatically calculated plaque burden as (plaque volume/vessel volume) × 100, where plaque volume was defined as the difference between the vessel and lumen volumes. VH-IVUS was used to characterize plaque composition into fibrous, fibro-fatty, necrotic core, and dense calcium components. The tissue component volumes and percentages, including necrotic core volume and percentage, were automatically derived from VH-IVUS analysis (**Figure 2**). Plaque burden greater than 35% was classified as cases, and less than 35% as controls. In cases of multivessel disease, the mean plaque burden was used for plaque burden, and the mean necrotic core (NC) volume derived from VH-IVUS was used for necrotic volume. VH data were unavailable for 6 participants.

**Figure 2:**
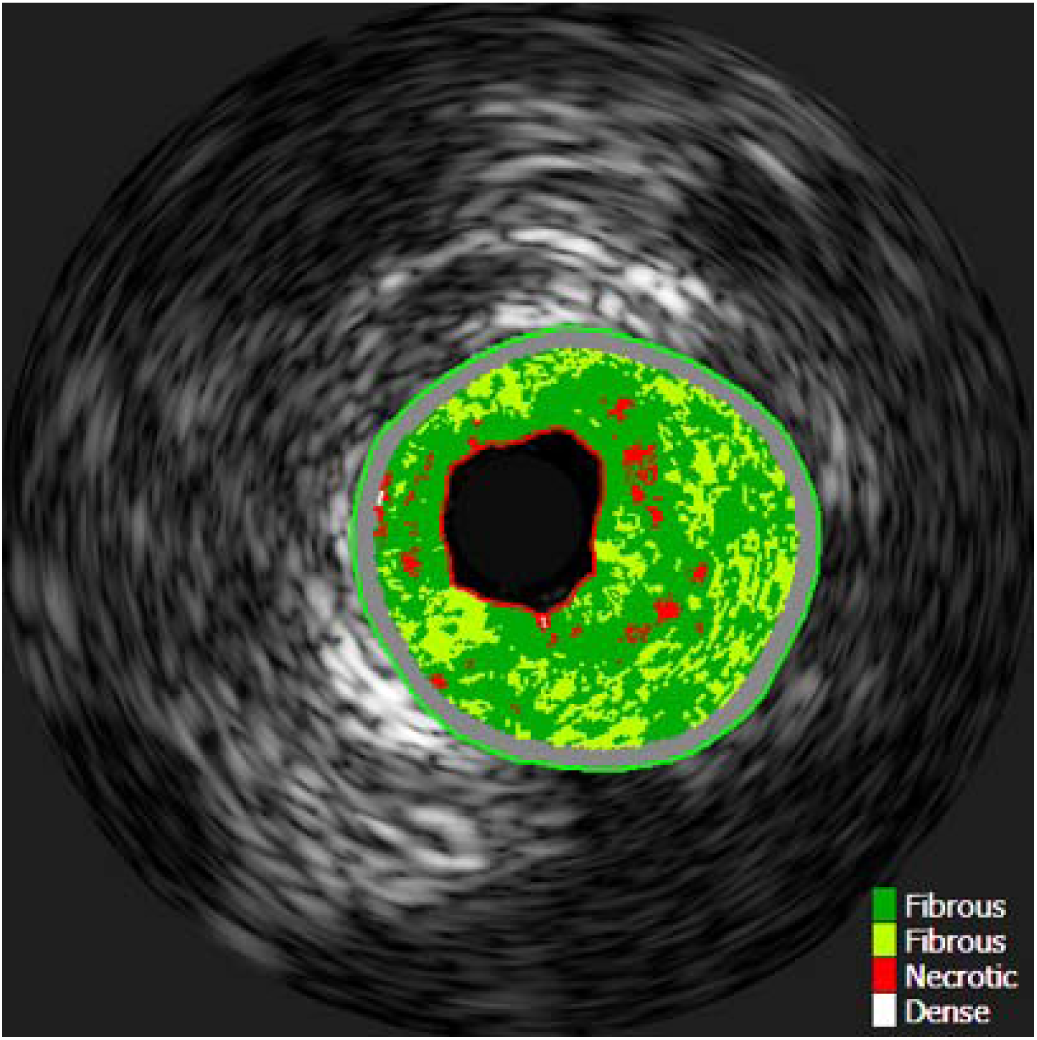
VH-IVUS characterization of coronary plaque composition. Color-coded plaque components identified by VH-IVUS include fibrous tissue (green), fibro-fatty tissue (light green/yellow), necrotic core (red), and dense calcium (white).

### PBMC isolation from whole blood

Blood samples were collected from all participants, and PBMC were isolated immediately using density gradient centrifugation. Briefly, 7ml of blood was collected and mixed with phosphate-buffered saline (Dulbecco’s) in a 1:1 ratio, carefully layered onto lymphocyte separation medium (LymphoSep TM, LSM), and then centrifuged at 5000 rpm for 30 minutes at room temperature. PBMCs isolated from the Buffy coat/plasma interface were washed with phosphate-buffered saline at 4 °C, flash frozen, and stored at –80 °C till further use.

### RNA Isolation from PBMCs

RNA was extracted from PBMCs using the mirVana™ miRNA Isolation Kit (AM1560, AM1561) according to the manufacturer’s instructions. Briefly, PBMCs were lysed with lysis buffer and subjected to acid-phenol-chloroform extraction. Following centrifugation, the upper aqueous phase was collected, precipitated with 100% ethanol, and subjected to centrifugation. The resulting RNA pellet was washed, resuspended, filtered through filter cartridges, quantified with a Qubit fluorometer, and stored at –80 °C until further use.

### Small RNA sequencing & bioinformatics analysis

Small RNA sequencing was performed using either the Illumina TruSeq Small RNA Library Preparation protocol or the QIAseq miRNA Library Kit. Both platforms follow a similar workflow. Briefly, for library preparation, 100 ng of RNA was ligated to 3′ and 5′ adapters, followed by reverse transcription and PCR amplification to generate cDNA libraries, during which unique molecular indices (UMIs) were incorporated. The resulting cDNA was purified using magnetic beads and amplified by PCR using compatible indexing primers. The amplified libraries were subsequently cleaned with magnetic beads to generate the final miRNA sequencing libraries. Library quality and concentration were assessed using the Agilent TapeStation D1000 ScreenTape (5067–5582) and the Invitrogen Qubit 4 High Sensitivity (HS) DNA Kit (Q33231). The prepared libraries were pooled and sequenced on an Illumina HiSeq 2500 or NovaSeq X Plus platform using 150 × 2 paired-end chemistry.

Raw sequence reads were assessed for quality using FastQC, and adapter sequences as well as low-quality reads were trimmed using Cutadapt (v3.5). Clean, high-quality reads were subsequently processed with the miRDeep2 (v0.1.3) pipeline using the miRbase and human reference genome GRCh38/hg38 to quantify the miR counts. Differential expression analysis was performed using DESeq2. miRs with very low expression were filtered out, and miRs with an absolute log2 fold change (|log2FC|) ≥ 0.9 and a p-value < 0.05 were considered to be significantly and differentially expressed. Normalized counts were calculated using a variance stabilizing transformation (VST) for correlation analysis with plaque burden and necrotic core (NC).

### Enrichment Analysis

Potential target genes of the selected miR were identified using three independent databases: TargetScan (https://www.targetscan.org/vert_80/), miRTarBase (https://mirtarbase.cuhk.edu.cn/∼miRTarBase/miRTarBase_2025), and miRDB (https://mirdb.org/). TargetScan predictions were filtered based on a cumulative weighted context^++^ score ≤ −0.2 and the presence of strong seed matches (8mer, 7mer-m8, or 7mer-A1), resulting in 22 predicted targets. From miRTarBase, 88 experimentally validated target genes were retrieved, while 404 predicted targets were obtained from miRDB using the recommended score threshold (≥80). The combined gene list from all three databases was compiled and subjected to functional enrichment analysis using the Enrichr (https://maayanlab.cloud/Enrichr/) platform. Enrichment analysis was performed using the KEGG pathway database, and pathways with p-values < 0.05 were considered statistically significant.

### Statistical Analysis

All statistical analyses were performed using the R programming environment (version 4.5.2). Continuous variables are presented as mean ± standard deviation (SD). Intergroup differences were assessed using the Student’s *t-test* or the nonparametric Mann–Whitney U test after evaluation of normality using the Shapiro-Wilk Test. Categorical variables were compared using the chi-square test or Fisher’s exact test, as appropriate. Correlations between variables were determined using Pearson or Spearman rank correlation coefficients, depending on the results of the normality test. The diagnostic performance of miRs was evaluated using receiver operating characteristic (ROC) curve analysis, with area under the curve (AUC), sensitivity, and specificity reported. A two-tailed *t-test,* P< 0.05, was considered statistically significant.

## Results

### Demographic data of study participants

Two independent cohorts comprising participants from Mumbai (city) (Site 1) and Rishikesh (rural community) (Site 2). The baseline demographic and clinical characteristics of the Site 1 cohort are presented in **Table 1**. The Site 1 cohort included 24 participants, comprising 19 CAD (Cases) and 5 non-CAD (Controls) patients. There was no significant difference in mean age between the CAD and non-CAD patient cohorts. The prevalence of diabetes was higher in CAD patients than in non-CAD patients, but did not reach statistical significance, and the occurrence of hypertension was similar between the CAD and non-CAD patient cohorts **[Table 1]**.

**Table 1:** Data from Site 1 presented as mean ± SD for continuous variables and as percentages for categorical variables. Group comparisons were performed using appropriate statistical tests, and a p-value < 0.05 was considered statistically significant.

| Parameters | CAD (n = 19) | Non-CAD (n = 5) | p value |
| --- | --- | --- | --- |
| Age (years) | 53.79 ± 11.40 | 56.40 ± 8.08 | 0.638 |
| Diabetes (%) | 31.6 | 20.0 | 1.000 |
| Hypertension (%) | 63.2 | 60.0 | 1.000 |

The Site 2 cohort comprised 19 patients: 12 CAD cases and 7 non-CAD controls. The baseline demographic and clinical characteristics of this rural community cohort are presented in **Table 2**. The mean age was significantly higher in CAD patients than in non-CAD patients [**Table 2**]. Prevalence of diabetes was higher among CAD patients than among non-CAD participants [**Table 2**], but the difference did not reach statistical significance. However, hypertension was common in both groups, with no significant difference between the CAD and non-CAD groups [**Table 2**].

**Table 2:**
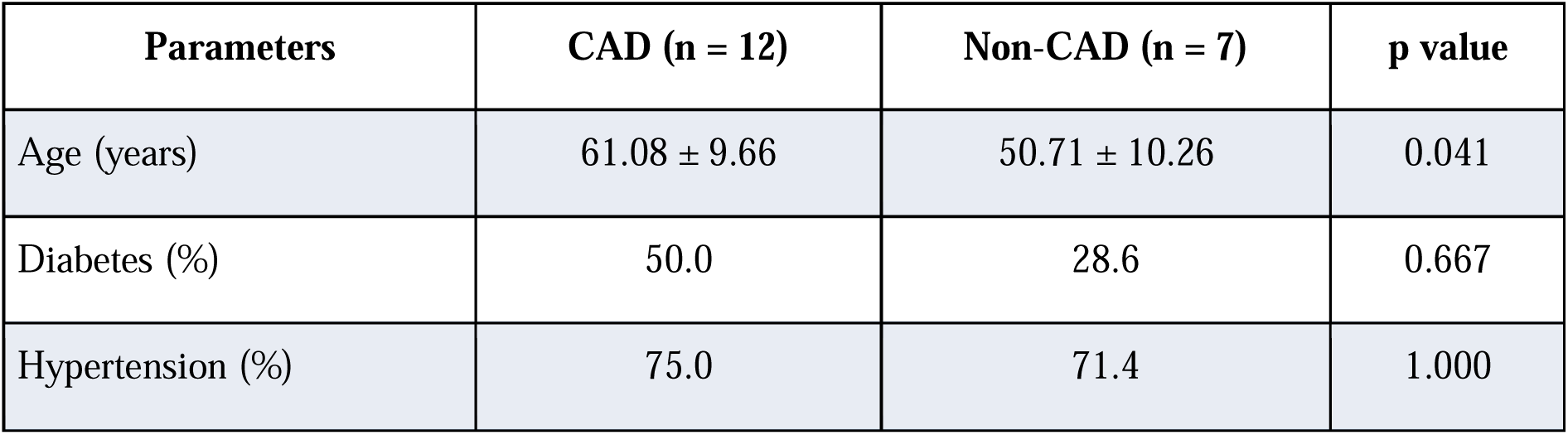
Data from Site 2 presented as mean ± SD for continuous variables and as percentages for categorical variables. Group comparisons were performed using appropriate statistical tests, and a p-value < 0.05 was considered statistically significant.

| Parameters | CAD (n = 12) | Non-CAD (n = 7) | p value |
| --- | --- | --- | --- |
| Age (years) | 61.08 ± 9.66 | 50.71 ± 10.26 | 0.041 |
| Diabetes (%) | 50.0 | 28.6 | 0.667 |
| Hypertension (%) | 75.0 | 71.4 | 1.000 |

### Differential miRNA expression in two independent populations

Unbiased global PBMC miR analysis of CAD and non-CAD patients from Site 1 revealed a subset of differentially expressed miR between these two groups. Differential expression analysis identified 2 upregulated and 13 downregulated miRs using the cutoff criteria outlined in the methods. These 14 altered miRs are listed in **Table 3** and the associated heatmap is shown in **Figure 3**.

**Figure 3:**
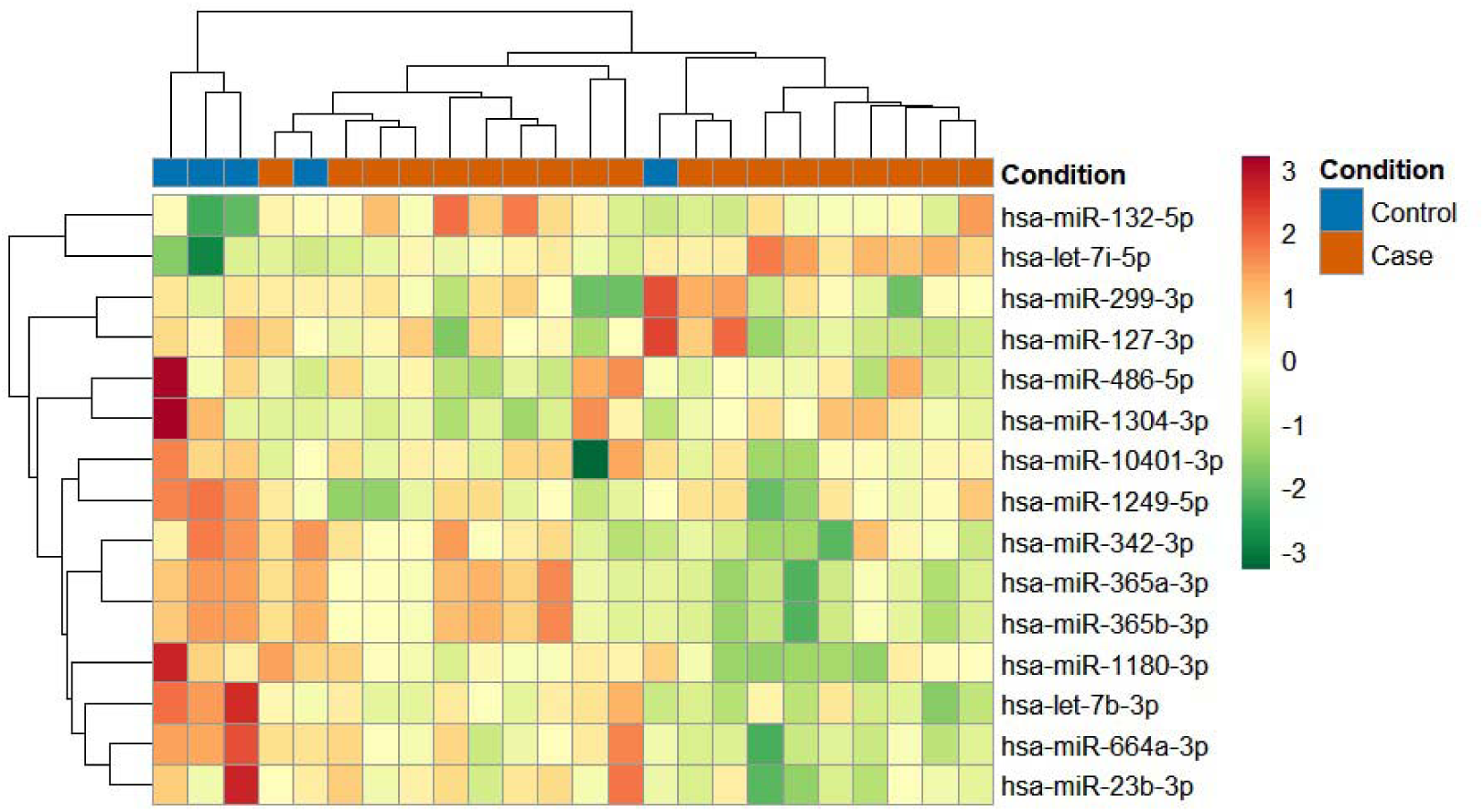
Heatmap of the top differentially expressed miRs from Site 1. The heatmap illustrates normalized expression levels of the most significantly dysregulated miRs across CAD (Cases) and non-CAD (Controls). Rows correspond to miRNAs and columns to individual samples.

**Table 3:** Top differentially expressed miRs in the Site 1 patient cohort showing log2 fold change (log2FC), base mean expression, and associated p values.

| miRNA | baseMean | log2FoldChange | p-value |
| --- | --- | --- | --- |
| hsa-let-7b-3p | 259.577 | -0.93496 | 0.00129 |
| hsa-let-7i-5p | 31372.25 | 0.819855 | 0.002493 |
| hsa-miR-10401-3p | 26.80481 | -1.01721 | 0.02535 |
| hsa-miR-1180-3p | 14.29706 | -1.12141 | 0.00327 |
| hsa-miR-1249-5p | 8.374793 | -1.00071 | 0.01157 |
| hsa-miR-127-3p | 429.9755 | -1.26645 | 0.02629 |
| hsa-miR-1304-3p | 606.3623 | -1.05981 | 0.01571 |
| hsa-miR-132-5p | 10.67649 | 0.856862 | 0.012829 |
| hsa-miR-23b-3p | 3243.964 | -0.70591 | 0.048757 |
| hsa-miR-299-3p | 11.0557 | -1.39969 | 0.04755 |
| hsa-miR-342-3p | 14554.37 | -0.73483 | 0.020682 |
| hsa-miR-365a-3p | 43.36038 | -0.82805 | 0.036275 |
| hsa-miR-365b-3p | 43.36038 | -0.82805 | 0.036275 |
| hsa-miR-486-5p | 345368.7 | -1.54859 | 0.00681 |
| **hsa-miR-664a-3p** | **583.6319** | **-1.01992** | **0.00327** |

Despite the variability associated with human studies, our heat map [**Figure 3**] shows a unique differential expression pattern of miRs, with marked separation of CAD (Cases) patients compared to non-CAD (Control) patients. This shows that unbiased PBMC miR expression analysis has potential as a non-invasive biomarker.

Interestingly, differential expression analysis of PBMC miR expression from Site 2 identified 23 upregulated and 20 downregulated miRs (n = 43) that met the cutoff criteria outlined in the methods. The top altered miRs are presented in **Table 4** with their differential expression reflected in the heat map [**Figure 4**].

**Figure 4:**
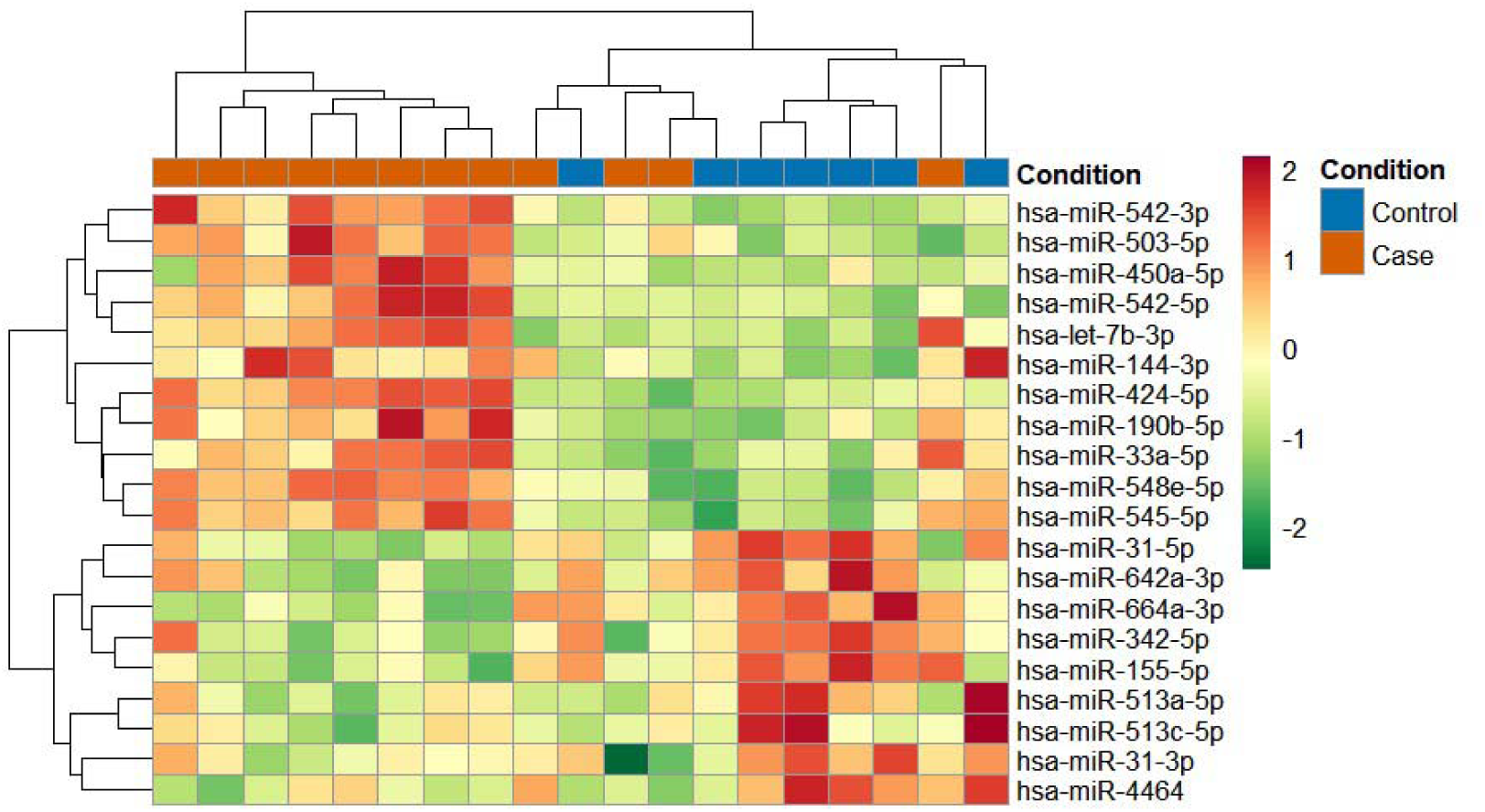
Heatmap of top differentially expressed miRs in CAD (Case) and non-CAD (Control) patient cohort from Site 2. Rows correspond to miRNAs and columns to individual samples.

**Table 4:**
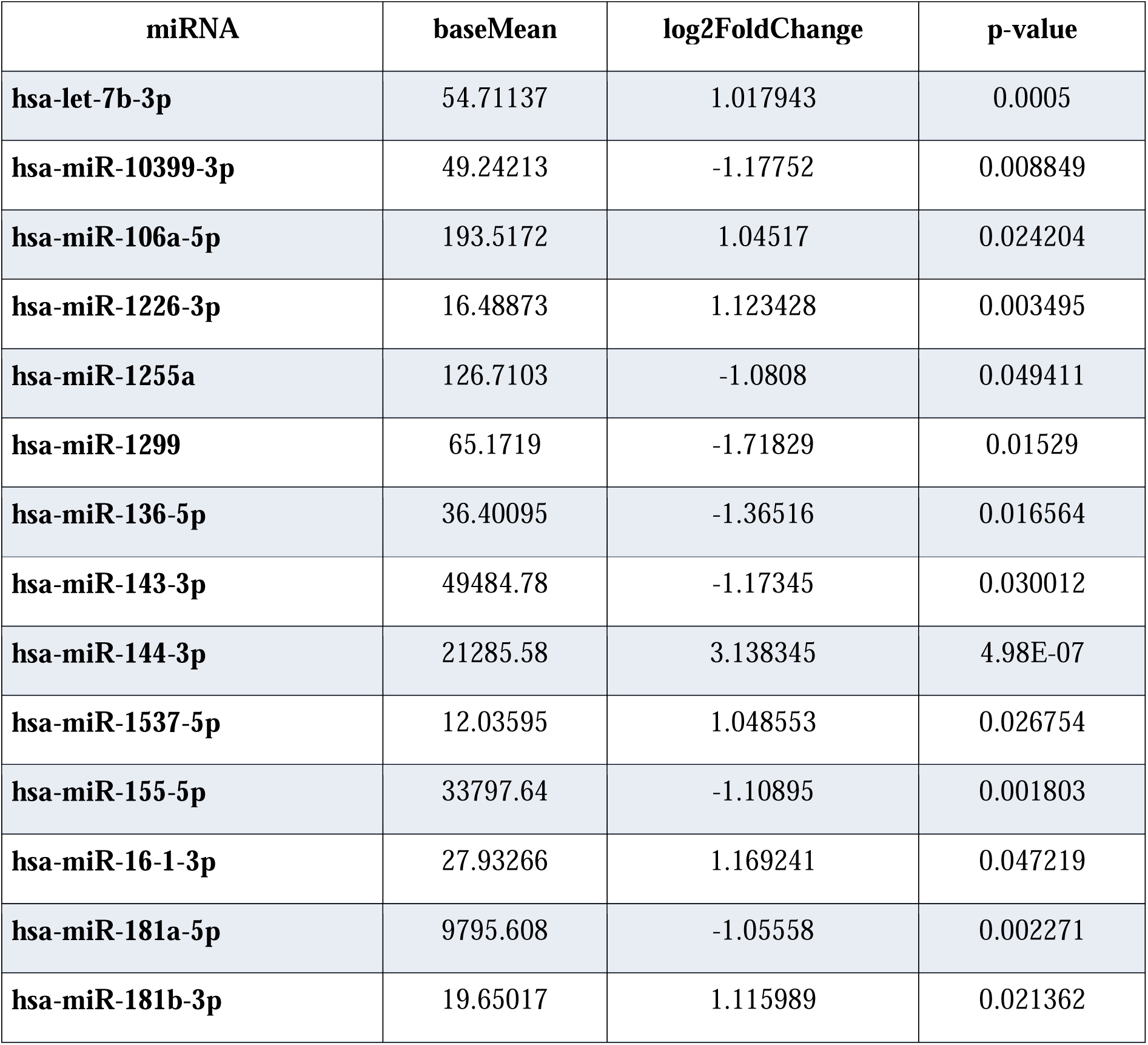
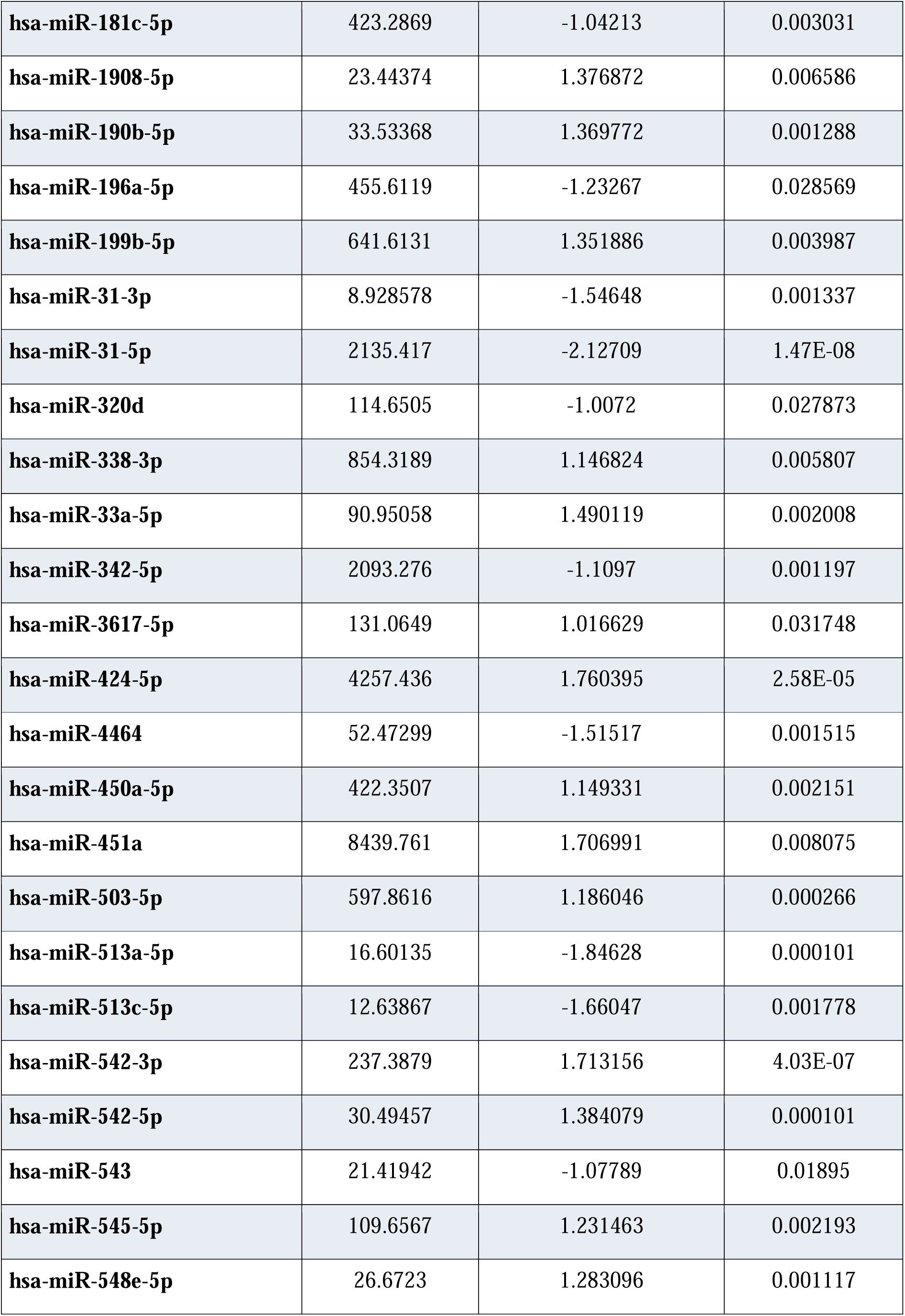
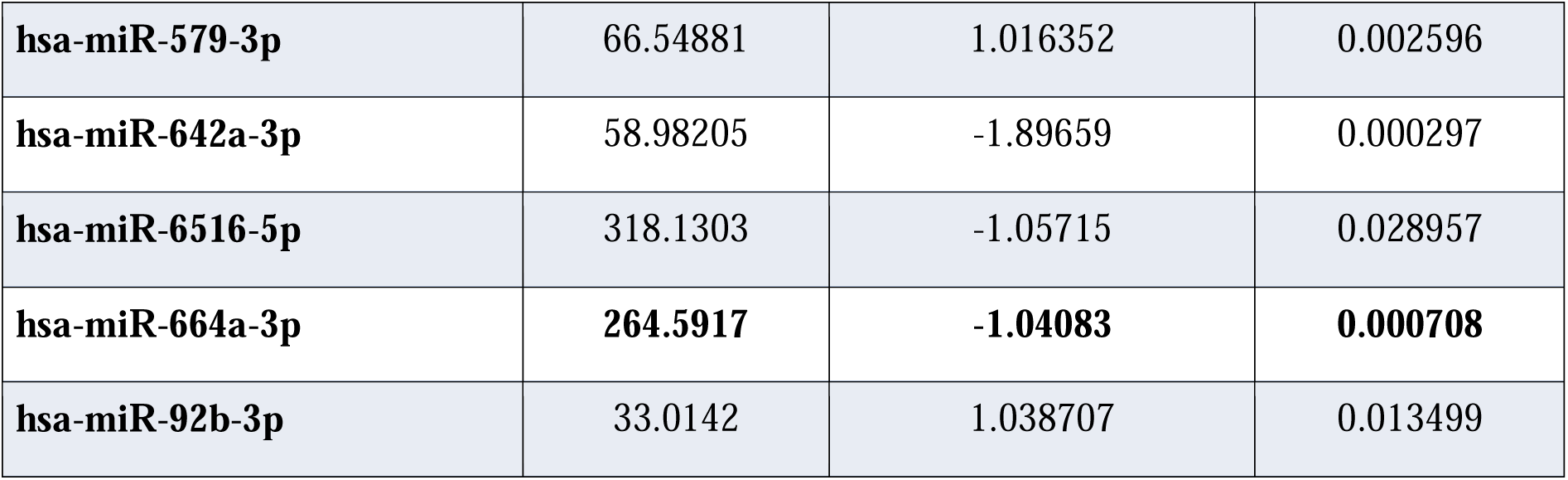
Top differentially expressed miRs in the Site 2 cohort, log2 fold change (log2FC), base mean expression, and associated p values.

### Common differentially expressed miRNAs across both independent populations

Since we observed that the number of significantly altered miRs are different between the sites, we evaluated whether there are common miRs between these two geographically distinct populations. Our comparative analysis shows that hsa-miR-664-3p is consistently downregulated in CAD patients across both patient cohorts compared to non-CAD patients. However, hsa-let-7b-3p exhibited discordant patterns, with hsa-let-7b-3p elevated in site 2 CAD patients compared to its downregulation in site 1 patients. These findings show that miR-664a-3p exhibits reproducible differential expression across sites.

### Correlation of Plaque Burden With miR-664a-3p

Although miR664a-3p is consistently and significantly altered in CAD patient PBMCs compared to the non-CAD patient cohort, a key translational question is whether the changes in miR-664a-3p expression in each patient correlate with plaque burden measurements. Assessment of plaque burden measurements to expression of mir-664a-3p shows inverse correlation with a correlation coefficient r = –0.484 [**Figure 5, left panel**]. This association was statistically significant (p = 0.0165), indicating that decreased miR-664a-3p expression is associated with greater plaque burden in CAD patients from site 1. Similarly, normalized miR-664a-3p expression at site 2 showed an inverse correlation with plaque burden (r = −0.584), which was statistically significant (p = 0.0086) [**Figure 5, right panel**]. These findings show the potential of miR-664a-3p as a surrogate marker for plaque characteristics in CAD patients.

**Figure 5.**
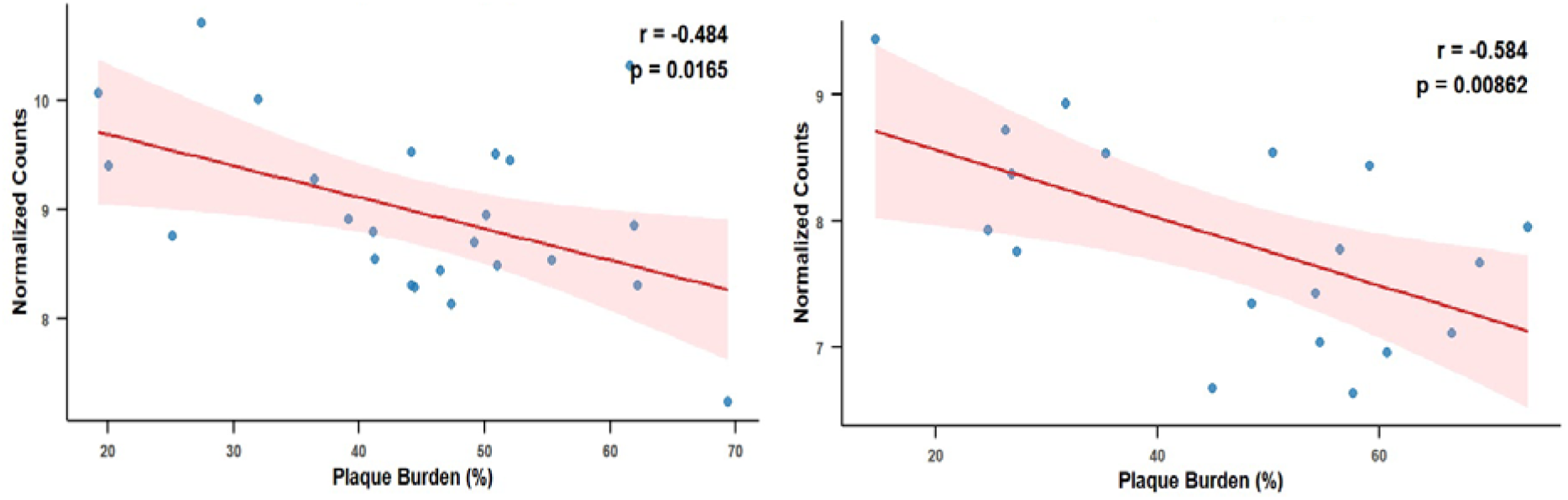
Pearson correlation of normalized miR-664a-3p expression with % plaque burden in the patient cohorts from site 1 (left panel) and site 2 (right panel).

### Correlation of miR-664a-3p Expression With Necrotic Core Volume

Since miR-664a-3p showed an inverse correlation with % plaque burden, we tested whether this association also holds for necrotic core volume, which in part determines plaque stability. Pearson correlation between normalized miR-664a-3p expression for each patient and their necrotic core volume at Site 1 showed an inverse correlation (r = –0.429) [**Figure 6, left panel**]. Furthermore, this correlation was statistically significant, p = 0.0478, [**Figure 6, left panel**]. Similarly, correlation analysis from site 2 showed significant inverse correlation with necrotic core volume (r = –0.578, p = 0.0239) [**Figure 6, right panel**]. Together, these studies show that miR664a-3p can serve as a non-invasive surrogate for assessing plaque characteristics in CAD, with the potential for direct correlation with plaque burden progression.

**Figure 6.**
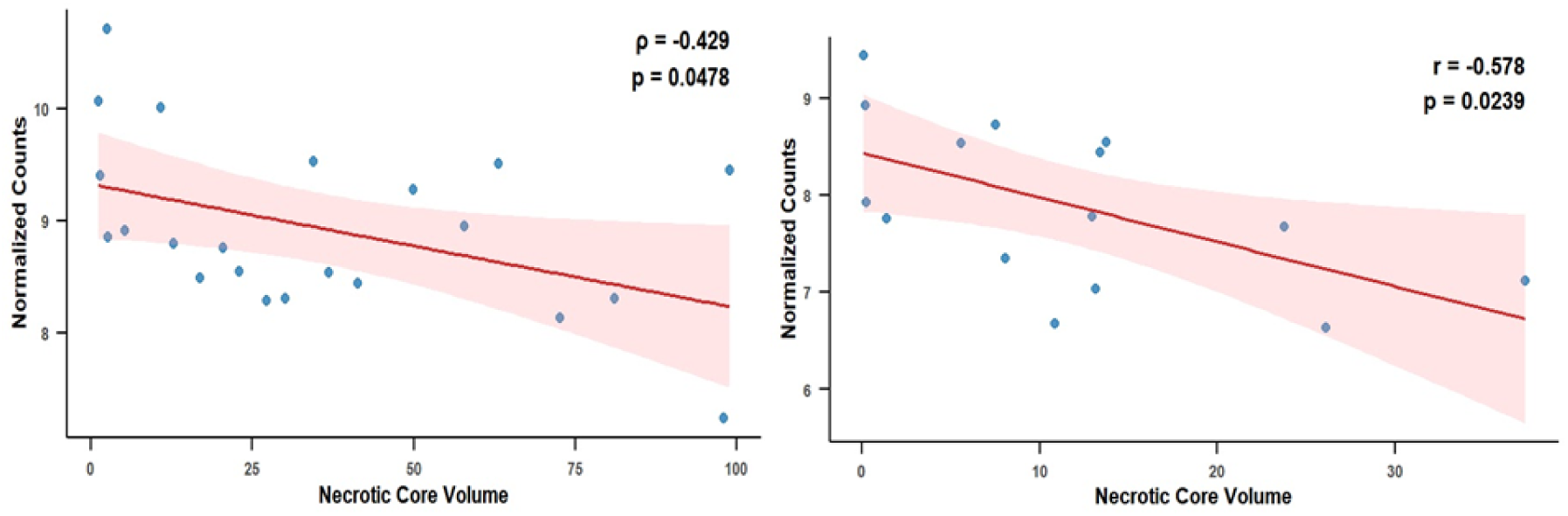
Pearson correlation of normalized miR-664a-3p expression with necrotic core volume in patient cohorts from site 1 (left panel) and site 2 (right panel).

### Diagnostic Performance of miR-664a-3p

To assess the diagnostic value and rigor of miR664a-3p in relation to plaque characteristics, a Receiver Operating Characteristic (ROC) curve analysis was performed. Using the Site 1 (Mumbai) cohort of 24 patients for the ROC analysis showed that the area under the curve (AUC) was 0.842 (CI: 0.647-1) [**Figure 7, left panel**], while the independent Site 2 (Rishikesh) cohort of 19 patients showed that the AUC was 0.881 (CI:0.73-1) [**Figure 7, right panel**]. These data show consistent and reproducible discriminatory performance across both cohorts for assessing the diagnostic value of miR-664a-3p as a potential non-invasive biomarker in PBMCs from CAD patients, with an innate ability to be linked to plaque burden.

**Figure 7.**
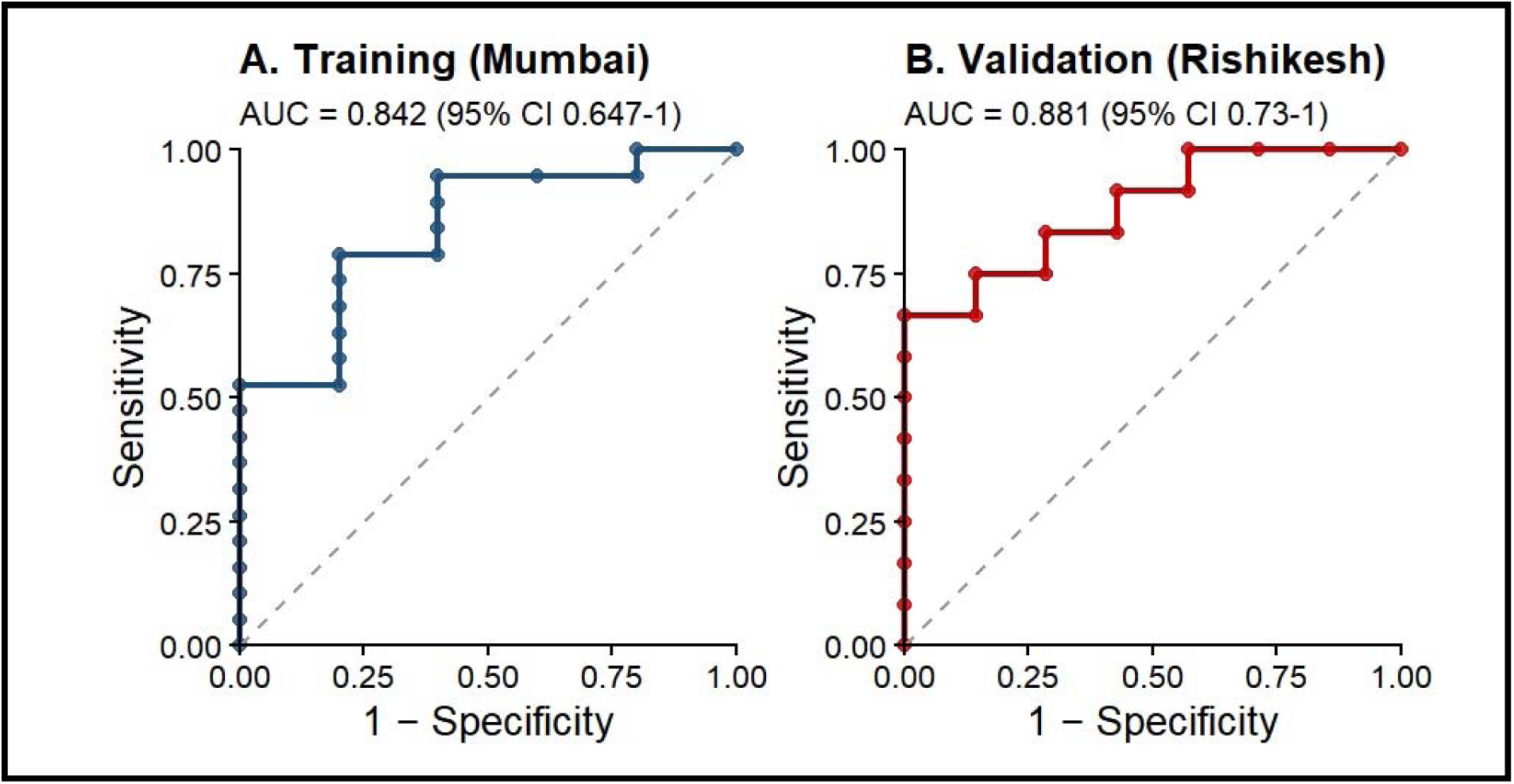
ROC analysis of miR-664a-3p for CAD with ROC curves showing the diagnostic performance of miR-664a-3p in the site 1 (Mumbai) cohort. (**left panel, A**) and site 2 (Rishikesh) cohort (**right panel, B**). The AUC with 95% confidence intervals is depicted for each cohort from each site.

## Discussion

Our findings show that PBMC miR-664a-3p could serve as a robust and reproducible biomarker significantly associated with both total plaque burden and necrotic core characteristics in an Indian patient population with CAD from two distinct geographical locations. The results show a consistent reduction in miR-664a-3p across two independent Indian patient cohorts, despite a significant age difference between CAD and non-CAD patients at site 2. Furthermore, miR-664a-3p is inversely correlated with plaque characteristics in each patient, reflecting its strong predictive capability. Consistently, high fidelity and comparable ROC analysis of miR-664a-3p across different geographical sites show promising translational potential for using PBMC miR-664a-3p as a non-invasive surrogate marker for assessing plaque characteristics, including instability and risk stratification, in the Indian population with CAD.

Several studies have reported the potential of miR-126 as a biomarker that is downregulated in patients with CAD and its association with advanced or unstable plaque phenotypes (14,15). miR-126 is considered to originate from endothelial cells due to its enrichment in endothelial cells. miR-126 is thought to be critical in maintaining vascular integrity, regulating angiogenic signaling, and limiting inflammatory activation by suppressing VCAM-1 expression (16). Its reduced expression has been directly linked to endothelial dysfunction and accelerated atherogenesis (16, 17). The observed inverse relationship between miR-126 levels and plaque burden supports the idea that loss of protective miR-126 may lead to upregulation and stabilization of deleterious proteins that could contribute towards adverse vessel remodeling and lesion progression (14,15).

Endothelial cells are considered a potential source of serum miR-126 due to their enrichment in these cells, but it remains challenging to state with certainty that serum miR-126 in CAD patients originates from endothelial cells. Given the dynamic regulation of miR-126, the origin of serum miR-126 could be attributed to structural cells, such as smooth muscle cells, or inflammatory cells, which also respond to changes in vessel plaque architecture. To overcome this uncertainty regarding the origin of the miRs, we evaluated the miR signature in PBMCs from CAD versus non-CAD patients and correlated it with plaque features measured by IVUS, the gold standard for evaluating plaque characteristics. Since PBMCs act as sentinels and mediate a reparative inflammatory response to vessel injury or changes in vessel homeostasis, we hypothesized that dynamic miR fluctuations in PBMCs would reflect ongoing alterations in plaque architecture. Also, PBMCs are recognized as sensitive indicators of systemic immune dysfunction and inflammatory milieu in CAD (18,19). Therefore, the link between PBMC miRs and necrotic core expansion that occurs with the progression of CAD reflects an underlying changing inflammatory landscape that is central to the transition from stable to vulnerable plaques (16). Unbiased small RNA sequencing shows that PBMC miR-664a-3p is the most significantly downregulated miR in PBMCs and that it tracks remarkably well with plaque characteristics, including necrotic core volume. However, in our studies, miR-126 in PBMCs was not significantly altered in CAD patients compared with non-CAD participants. This suggests that PBMC miR-664a-3p can serve as a viable non-invasive biomarker because its changes inversely correlate with plaque characteristics with high fidelity in two geographically distinct cohorts of the Indian population.

miR-664a-3p has been characterized in malignancies as its dysregulation and oncogenic role have been documented in gastric, lung, and pancreatic cancers (20,21). These studies show that elevated levels of miR-664a-3p in cancer cells are a key driver of cell proliferation (20,21). Similarly, altered expression of serum miR-664a-3p has also been reported in inflammatory conditions such as chronic periodontitis and in cardioembolic stroke, suggesting its broader involvement in systemic inflammatory or vascular signaling pathways (22,23). Similarly, serum miR profiling of patients with obstructive sleep apnea showed significant downregulation of miR-664a-3p. Interestingly, the reduction in serum miR-664a-3p in patients with obstructive sleep apnea was also associated with increased carotid intima-media thickness on ultrasonography. This suggests that miR-664a-3p can be used as a potential diagnostic tool for atherosclerosis in patients with obstructive sleep apnea, as obstructive sleep apnea is known to be associated with a higher incidence of atherosclerosis (16,25). As mentioned, a limitation is the cellular origin of miR-664a-3p. This is critical because changes in secreted miR may not originate from the diseased organ but could arise from secondary sources and may not directly reflect the progression of pathology with high fidelity.

The robust correlation of miR-664a-3p with plaque burden and necrotic core characteristics across two geographically diverse patient cohorts suggests that it may serve as a novel surrogate diagnostic molecular marker for the Indian population. Such a rationale is supported by the observation that PBMC miR-664a-3p shows an inverse correlation with plaque characteristics, specifically necrotic core volume, measured by IVUS imaging. This is further supported by the high confidence interval observed in the ROC analysis and the area under the curve, which reflects the significant diagnostic performance of PBMC miR-664a-3p in vascular remodeling with plaque pathology. We believe the unique strength of our study in determining changes in the miR profile in PBMCs gives us a window into understanding whether PBMC cellular responses reflect the progressive pro-inflammatory changes that occur in vascular plaque pathology. Consistent with this idea, functional enrichment analysis of the predicted and validated gene targets of miR-664a-3p reveals regulation of pathways key to inflammatory responses, vascular signaling, and cellular stress mechanisms.

The enrichment of IL-17 and TNF signaling pathways suggests that miR-664a-3p may negatively regulate genes involved in inflammatory signaling networks and vascular homeostasis. Thus, a reduction in miR-664a-3p would increase the expression of proteins in this pathway, leading to pro-inflammatory responses. Both IL-17 and TNF signaling are established mediators of chronic inflammation that facilitate the progression of atherosclerotic lesions by inducing endothelial dysfunction and promoting the feed-forward recruitment of immune cells (26). Specifically, IL-17 is detectable within atherosclerotic lesions and appears to exacerbate local tissue inflammation by stimulating the production of pro-inflammatory chemokines and cytokines, such as IL-6 and CXCL8, which drive monocyte and neutrophil infiltration (26,27). On the same note, chronic TNFα secretion by PBMCs may upregulate adhesion molecules, such as VCAM-1 and ICAM-1, and contribute to the expansion of the necrotic core, a hallmark of plaque vulnerability (28). These analyses indicate that targets of miR-664a-3p may contribute to inflammatory and vascular processes relevant to CAD. Therefore, dynamic regulation of miR-664a-3p in PBMCs could reflect ongoing changes in plaque pathology and could serve as a high-fidelity, non-invasive diagnostic tool that allows prediction of plaque stability and vulnerability in CAD patients.

## Limitations & Future Prospects

The modest sample size is a limitation; however, the significance observed in two geographically distinct Indian population cohorts provides a strong rationale for expanding and validating this approach using a multicenter population across India to establish miR-664a-3p as a robust biomarker for CAD. Furthermore, functional validation of miR-664a-3p would clarify its mechanistic role, unlocking the proinflammatory pathways.

## Conclusion

The three key features of this study are: 1) the identification of a PBMC miR-663a-3p as a PBMC signature, 2) the consistent downregulation of miR-664a-3p across two independent and geographically distinct Indian patient cohorts, and 3) the significant association of miR-664a-3p with both total plaque burden and necrotic core characteristics. Together, these findings position miR-664a-3p as a robust, non-invasive potential biomarker for CAD in the Indian population and for assessing plaque vulnerability. It is important as Indians are known to have a higher risk and rates of CAD compared to any ethnicity in the world.

## Funding

This work was supported by the Wellcome Trust–DBT India Alliance, the Wadhwani Foundation, intramural research funding from AIIMS Rishikesh, and iHub Drishti Foundation, IIT Jodhpur.

## Data Availability

All data produced in the present study are available upon reasonable request to the authors.

## Acknowledgements

The authors sincerely acknowledge the study participants for their valuable contribution to this research.

## Conflict of Interest

The authors declare no conflicts of interest related to this study.

## Notes

### Competing Interest Statement

The authors have declared no competing interest.

### Author Declarations

The institutional ethics committees of Grant Medical College & Sir JJ Group of Hospitals, Mumbai, and the All India Institute of Medical Sciences, Rishikesh, India, gave ethical approval for this work.

